# A Scalable Framework for Benchmarking Embedding Models for Semantic Medical Tasks

**DOI:** 10.1101/2024.08.14.24312010

**Authors:** Shelly Soffer, Benjamin S Glicksberg, Patricia Kovatch, Orly Efros, Robert Freeman, Alexander W Charney, Girish N Nadkarni, Eyal Klang

**Affiliations:** Institute of Hematology, Davidoff Cancer Center, Rabin Medical Center; Petah-Tikva, Israel; Charles Bronfman Institute of Personalized Medicine, Icahn School of Medicine at Mount Sinai, 1 Gustave L. Levy Place; New York, NY 10029, United States; Division of Data Driven and Digital Medicine, Department of Medicine, Icahn School of Medicine at Mount Sinai; New York, NY 10019, United States; Department of Genetics and Genomic Sciences, Icahn School of Medicine at Mount Sinai, New York, NY 10029, United States; School of Medicine, Tel Aviv University; Tel Aviv, Israel; National Hemophilia Center and Thrombosis Institute, Sheba Medical Center; Ramat Gan, Israel

## Abstract

Text embeddings convert textual information into numerical representations, enabling machines to perform semantic tasks like information retrieval. Despite its potential, the application of text embeddings in healthcare is underexplored in part due to a lack of benchmarking studies using biomedical data. This study provides a flexible framework for benchmarking embedding models to identify those most effective for healthcare-related semantic tasks. We selected thirty embedding models from the multilingual text embedding benchmarks (MTEB) Hugging Face resource, of various parameter sizes and architectures. Models were tested with real-world semantic retrieval medical tasks on (1) PubMed abstracts, (2) synthetic Electronic Health Records (EHRs) generated by the Llama-3-70b model, (3) real-world patient data from the Mount Sinai Health System, and the (4) MIMIC IV database. Tasks were split into ‘Short Tasks’, involving brief text pair interactions such as triage notes and chief complaints, and ‘Long Tasks’, which required processing extended documentation such as progress notes and history & physical notes. We assessed models by correlating their performance with data integrity levels, ranging from 0% (fully mismatched pairs) to 100% (perfectly matched pairs), using Spearman correlation. Additionally, we examined correlations between the average Spearman scores across tasks and two MTEB leaderboard benchmarks: the overall recorded average and the average Semantic Textual Similarity (STS) score. We evaluated 30 embedding models across seven clinical tasks (each involving 2,000 text pairs), across five levels of data integrity, totaling 2.1 million comparisons. Some models performed consistently well, while models based on Mistral-7b excelled in long-context tasks. ‘NV-Embed-v1,’ despite being top performer in short tasks, did not perform as well in long tasks. Our average task performance score (ATPS) correlated better with the MTEB STS score (0.73) than with MTEB average score (0.67). The suggested framework is flexible, scalable and resistant to the risk of models’ overfitting on published benchmarks. Adopting this method can improve embedding technologies in healthcare.

## INTRODUCTION

Text embeddings are numerical representations of text that capture the semantic meaning of words, phrases, or entire documents in a continuous vector space^1^. Currently, most text embeddings are generated by dedicatedly trained large language models (LLMs).

Text embeddings enable key tasks like semantic search and Retrieval Augmented Generation (RAG), which have transformative potential in various fields, including healthcare. ^2, 3^ Despite their potential, such advanced natural language processing (NLP) methods remain largely untapped in the medical domain. Traditional approaches to handling medical text often fall short in capturing the nuanced and specialized language used in clinical settings. This gap emphasizes the necessity for robust text embedding models that can handle the complexity and diversity of medical texts.

The Multilingual Text Embeddings Benchmark (MTEB), a known general case embedding models benchmarking framework, is designed to evaluate text embedding models across multiple domains ^4^. By providing standardized datasets and evaluation metrics, MTEB facilitates direct comparisons of model performance on tasks such as classification, clustering, and semantic textual similarity (STS). MTEB includes a broad array of datasets; however, its representation of the medical field remains limited, with datasets such as MedrxivClusteringP2P^5^, NFCorpus ^6^, and BIOSSES^7^ being small in size. This hampers the effective comparison of the latest embedding models with healthcare-specific text. Moreover, another well-known risk of existing benchmarks for LLMs is the risk of models’ overfitting on the published benchmark datasets.

This study aims to provide a flexible framework for evaluating the performance of leading text embedding models in capturing semantic similarity within medical texts. We used this framework to evaluate multiple embedding LLMs on multiple clinical and biomedical tasks.

## METHODS

### Overall Design

We aimed to identify and rank embedding models for semantic applications specific to the medical field. We selected thirty embedding models across different size groups from the MTEB leaderboard, available on GitHub ^8^, chosen based on their relevance and demonstrated performance metrics. We designed a series of tasks simulating real-life medical semantic retrieval challenges to test models’ performance in embedding medical terminology and contextual nuances. To evaluate the performance under varying data quality conditions, we designed an experiment where we deliberately introduced noise into the text pairs used. We then used Spearman rank correlation analysis to measure the relationship between the models’ performance and the different levels of data integrity we created. This research was conducted with the approval of the Institutional Review Board (IRB) of the Mount Sinai Health System.

### Model Selection Criteria

Our study evaluated models listed on the MTEB leaderboard. We selected open-source models available on Hugging Face with implementations via the sentence transformers library. Each model’s inclusion required the presence of implementation code within its Hugging Face model card. We systematically chose five models from each of MTEB size groups: ’<0.1 billion parameters’, ‘0.1-0.25 billion parameters’’, ‘0.25-0.5 billion parameters’’, ‘0.5-1 billion parameters’’, ‘1-5 billion parameters’’, and ‘>5 billion parameters’’, prioritizing the best-performing models in each category. Among these, ‘Bio_ClinicalBERT’ served as a baseline reference ^9^. This model utilizes contextual embeddings derived from Google’s BERT architecture and is trained on PubMed and the MIMIC III dataset. Notably, ‘Bio_ClinicalBERT’ was not specifically trained for semantic embedding tasks, making it a standard benchmark against more specialized models. All included evaluated models and their characteristics are detailed in **Table 1**.

**Table 1:** Model Details According to Model Size.

| Model size | Model | Memory Usage (GB, fp32) | Window Size (Tokens) | Model Size (Million Parameters) |
| --- | --- | --- | --- | --- |
| <0.1GB | GIST-small-Embedding-v0 <sup>11</sup> | 0.12 | 512 | 33 |
|  | bge-small-en-v1.5 <sup>19</sup> | 0.12 | 512 | 33 |
|  | gte-small <sup>20</sup> | 0.12 | 512 | 33 |
|  | all-MiniLM-L12-v2 <sup>21</sup> | 0.12 | 512 | 33 |
|  | all-MiniLM-L6-v2 <sup>22</sup> | 0.09 | 512 | 23 |
| 0.1-0.25GB | GIST-Embedding-v0 <sup>11</sup> | 0.41 | 512 | 109 |
|  | all-mpnet-base-v2 <sup>23</sup> | 0.41 | 512 | 110 |
|  | gte-base <sup>20</sup> | 0.41 | 512 | 109 |
|  | e5-base-4k <sup>24</sup> | 0.42 | 4096 | 112 |
|  | Bio_ClinicalBERT <sup>9</sup> | 0.41 | 512 | 110 |
| 0.25-0.5GB | mxbai-embed-large-v1<sup>25</sup> | 1.25 | 512 | 335 |
|  | UAE-Large-V1<sup>25</sup> | 1.25 | 512 | 335 |
|  | GIST-large-Embedding-v0<sup>11</sup> | 1.25 | 512 | 335 |
|  | bge-large-en-v1.5<sup>19</sup> | 1.25 | 512 | 335 |
|  | b1ade-embed<sup>12</sup> | 1.25 | 512 | 335 |
| 0.5-1GB | multilingual-e5-large-instruct<sup>26</sup> | 2.09 | 514 | 560 |
|  | multilingual-e5-large<sup>26</sup> | 2.09 | 514 | 560 |
|  | Solon-embeddings-large-0.1 | 2.09 | 512 | 560 |
|  | bge-m3-custom-fr<sup>27</sup> | 2.12 | 8192 | 568 |
|  | mmlw-e5-large<sup>28</sup> | 2.09 | 514 | 560 |
| 1-5GB | instructor-xl<sup>29</sup> | 4.62 | 512 | 1241 |
|  | sentence-t5-xl<sup>30</sup> | 4.62 | 512 | 1241 |
|  | sentence-t5-xxl<sup>30</sup> | 18.12 | 512 | 4865 |
|  | gtr-t5-xxl <sup>31</sup> | 18.12 | 512 | 4865 |
|  | SGPT-2.7B-weightedmean-msmarco-specb-bitfit <sup>32</sup> | 10.0 | 2048 | 2685 |
| >5GB | SFR-Embedding-Mistral <sup>33</sup> | 26.49 | 32768 | 7111 |
|  | e5-mistral-7b-instruct <sup>34</sup> | 26.49 | 32768 | 7111 |
|  | Linq-Embed-Mistral <sup>35</sup> | 26.49 | 32768 | 7111 |
|  | NV-Embed-v1 <sup>13</sup> | 29.25 | 32768 | 7851 |
|  | SFR-Embedding-2_R <sup>36</sup> | 26.49 | 32768 | 7111 |

### Databases

We extracted medical data from the following databases

1. <u>PubMed</u>. Abstracts were extracted using the MeSH terms: “Artificial Intelligence”, “Machine Learning”, and “Deep Learning,” spanning the last five years. We ensured abstracts and keywords were non-null. Paired keywords were directly extracted from PubMed. Additionally, using Llama-3-70b, we generated search queries from a collection of PubMed abstracts (The complete prompt with the JSON directive and formatting is in **Supplementary eFigure 1**).
2. <u>LLM Synthetic Electronic Health Records (EHR) Notes</u>. Synthetic notes were generated using the Llama-3-70b model to create a simulated dataset of EHRs. The notes were based on three term list (**Supplementary eFigure 2)** to ensure variability in the notes. Using Llama-3-70b, we also generated search queries paired with the synthetic notes (The complete prompt with the JSON directive and formatting is in **Supplementary eFigure 3**).
3. <u>Mount Sinai Health System (MSHS) EHR</u>. Clinical care data from actual patients from 2023, including triage notes, chief complaints, physician notes and admission H&P notes.
4. <u>MIMIC IV Database.</u> This open database of de-identified medical information served as a source for chest X-ray reports and discharge notes. ^10^ Discharge notes were summarized using Llama-3-70b (The complete prompt with the JSON directive and formatting is in **Supplementary eFigure 4)**.

### Embedding Tasks Overview

**Table 2** outlines the data sources and configurations for our embedding tasks. Each task in our study involved 2,000 pairs of source and destination text. These pairs were organized into two categories: ‘Short Tasks’ for brief text interactions and ‘Long Tasks’ for more extended text analyses. Below, we detail each task type.

**Table 2:** Overview of embedding tasks. Abbreviations: LLM: Large Language Model, EHR: Electronic Health Record, MSHS: Mount Sinai Health System, CXR: Chest X-ray, ICU: Intensive Care Unit, ED: Emergency Department, H&P: History and Physical

| Length | Data Source | Source | Destination |
| --- | --- | --- | --- |
| Short Tasks | PubMed | Abstracts | Search queries |
|  | PubMed | Abstracts | Keywords |
|  | LLM Synthetic EHR Notes | Synthetic notes | Search queries |
|  | MSHS EHR | Triage Notes | Chief Complaints |
|  | MIMIC IV CXR Reports | Imaging Findings | Imaging Impressions |
| Long Tasks | MIMIC IV EHR Notes | ICU discharge notes | LLM Generated Summaries |
|  | MSHS EHR | ED Physician Notes | H&P Notes |

### Short Tasks

PubMed (Abstracts to Queries): To link abstracts with generated queries they inspired. PubMed (Abstracts to Keywords): To match abstracts with extracted keywords.

LLM Synthetic EHR Notes: To connect synthetic notes with corresponding search queries.

MSHS EHR: To pair triage notes with corresponding chief complaints.

MIMIC IV Chest-XR Reports: To link observations noted in the ‘Findings’ section with interpretations from the ‘Impression’ section. To ensure unique findings and impressions, we first filtered out non-significant (“normal”) impressions. A complete list of terms used to categorize “normal” findings is presented in **Supplementary eTable 1**.

### Long Tasks

MIMIC IV Discharge Notes: To match a random sample of discharge notes with their summaries generated using Llama-3-70b.

MSHS ED Physician Notes: To concatenate random sample of 2000 admitted patient cases from 2023, with corresponding admission H&P notes.

The selection criteria and process for data preparation, including the use of specific search parameters and data handling rules, including detailed prompts used for Llama-3-70b implementations are available in the **supplementary materials**.

### Experimental Setup

We evaluated the performance of embedding models across varying levels of data integrity. The experimental design incorporated a gradient of data integrity levels to simulate different degree of data alteration:

- **0% Integrity**: Utilized pairs with unmatched source and destination (X source - Y destination).
- **25% Integrity**: Combined 25% of the original source (X) with 75% of an alternate source (Z), paired with the original destination (X destination).
- **50% Integrity**: Mixed 50% of the original source (X) with 50% of an alternate source (Z), paired with the original destination (X destination).
- **75% Integrity**: Merged 75% of the original source (X) with 25% of an alternate source (Z), paired with the original destination (X destination).
- **100% Integrity**: Paired the original source with its corresponding original destination (X source - X destination)

For short tasks, we used the first % of the original source string and completed it with the remaining (100%) from an alternate source. For long tasks, 25% integrity meant each one sentence from the original source (1/4) was followed by three sentences from an alternate source (3/4); 50% data integrity meant two sentences from each source (2/4, 2/4), and so on. We used different techniques for adding noise because short tasks may not have enough sentences for the long method, and in long tasks, short context window models are truncated at 512 tokens, which would primarily capture the ‘X’ part if we used the short task method. Using this approach allowed us to conduct 10,000 comparisons per task (5x2000).

We used Spearman rank correlation to analyze the relationship between the level of data integrity and the performance metrics (cosine similarity, Euclidean difference, dot product). This methodology was designed to evaluate how effectively each model captures and maintains the key semantic features of the source material at different integrity levels. All computations and model evaluations were performed on a dedicated MSHS server equipped with H100 80GB GPUs. The analyses were conducted using Python version 3.9.18, with additional dependencies on several key libraries: PyTorch version 2.2.2+cu121, Transformers version 4.41.2, Sentence Transformers version 3.0.1, pandas version 2.1.4, and scikit-learn version 1.3.0.

### Statistical Analysis Methods

To evaluate the performance of embedding models, we applied cosine similarity, Euclidean difference, and dot product metrics to measure the similarity or distance between source and destination vectors. For each integrity level, we assessed the models using these three metrics. We then calculated the Spearman rank correlation between the integrity levels and the performance metrics to determine the robustness and reliability of the models across varying data quality levels.

We also examined correlations between the average Spearman correlation for each model across tasks and two established MTEB leaderboard scores: the model’s overall recorded average score and the average Semantic Textual Similarity (STS) score. Additionally, STS scores were correlated to average MTEB scores to contextualize our findings.

To ensure that the natural variance of Spearman scores across tasks did not skew the overall assessment of each model, we ranked the models independently for each type of task—short, long, and overall. We used the ‘Bio_ClinicalBERT’ model as a reference, assigning it a baseline rank of one for all tasks . Other models were then ranked according to how their Spearman scores compared to this reference (sorted order). We also compared model performance between clinical and PubMed tasks.

## RESULTS

### Data Overview

We assessed 30 embedding models across seven clinical embedding tasks, each involving 2,000 pairs of notes at different levels of integrity, from unchanged to fully mixed. This evaluation encompassed a total of 2.1 million comparisons, calculated as 30 models * 7 tasks * 5 levels of integrity * 2,000 pairs. The number of vectors was slightly higher, as the calculations also included destination vectors. This adjustment resulted in a total of 2.52 million vectors, computed as 30 models * 7 tasks * 6 (5 level of integrity and additional destination vector) * 2,000 pairs.

**Table 3** presents the variations in word counts for different clinical embedding tasks, categorized as “Short Tasks” and “Long Tasks.” These categories illustrate the range of textual demands, from concise interactions to more detailed clinical texts.

**Table 3:**
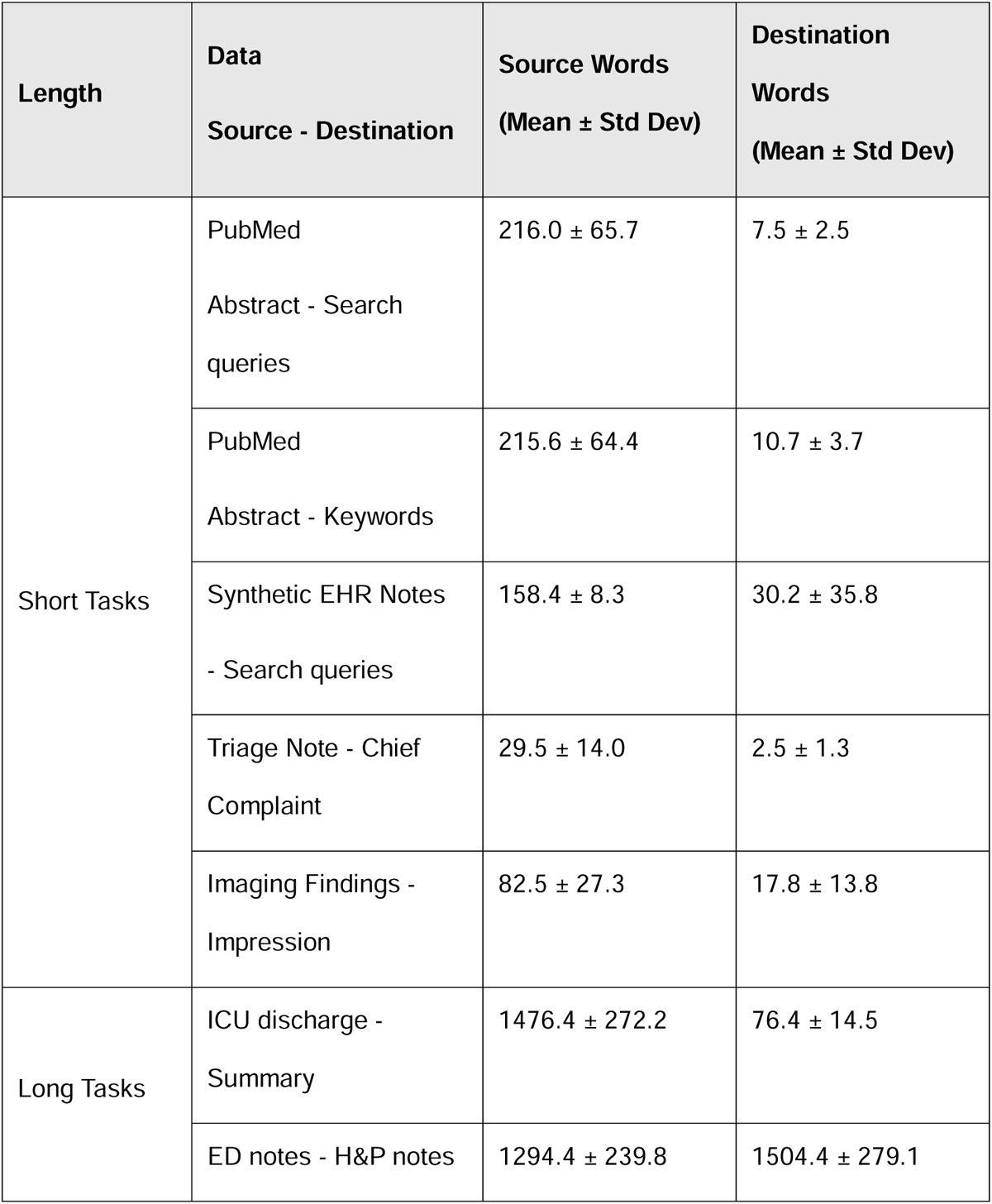
Lengths of tasks inputs. Abbreviations: EHR: Electronic Health Record, ICU: Intensive Care Unit, ED: Emergency Department, H&P: History and Physical

### Metric Efficacy Across Models and Tasks

We evaluated three metrics—cosine similarity, Euclidean difference, and dot product— across multiple embedding models and tasks to determine their efficacy in capturing semantic similarity under different integrity levels. We used Spearman rank correlation to evaluate how the metrics correlate with varying levels of data integrity, which reflect model performance. Cosine similarity emerged as the most effective metric overall in maintaining semantic integrity across integrity levels. Model-specific performances for the best-performing metric are available in **Supplementary Excel Table 4**. While most models showed minimal variation in metric efficacy between tasks, some differences were observed in specific cases, such as ‘e5-base-4k’ in the ‘imaging’ task, ‘mmlw-e5-large’ in the ‘PubMed query’ task, and ‘Bio_ClinicalBERT’ in the ‘QA’ task.

**Table 4:** Spearman Correlation Rankings and Average Task Performance Score (ATPS). This table presents the Spearman correlation rankings for each of the 30 embedding models across the seven clinical embedding tasks. Additionally, the table includes the ATPSs, which provide an average performance indicator for each model across all tasks. Abbreviations: ED: Emergency Department, H&P: History & Physical Notes, CXR: Chest X-ray, EHR: Electronic Health Record

| Model size | Model | Discharge Notes to Summaries | ED to H&P | Triage Notes to Chief Complaints | CXR Findings to Impressions | PubMed Abstracts to Keywords | PubMed Abstracts to Queries | Synthetic EHR to Search Queries | Average Task Performance Score (ATPS) |
| --- | --- | --- | --- | --- | --- | --- | --- | --- | --- |
| <0.1GB | GIST-small-Embedding-v0 <sup>11</sup> | 0.71 | 0.54 | 0.4 | 0.4 | 0.72 | 0.78 | 0.84 | 0.63 |
|  | bge-small-en-v1.5 <sup>19</sup> | 0.67 | 0.37 | 0.32 | 0.38 | 0.69 | 0.76 | 0.81 | 0.57 |
|  | gte-small <sup>20</sup> | 0.69 | 0.5 | 0.42 | 0.41 | 0.69 | 0.75 | 0.75 | 0.6 |
|  | all-MiniLM-L12-v2 <sup>21</sup> | 0.62 | 0.26 | 0.38 | 0.32 | 0.58 | 0.66 | 0.55 | 0.48 |
|  | all-MiniLM-L6-v2 <sup>22</sup> | 0.7 | 0.4 | 0.4 | 0.36 | 0.67 | 0.77 | 0.79 | 0.58 |
| 0.1-0.25GB | GIST-Embedding-v0 <sup>11</sup> | 0.72 | 0.52 | 0.45 | 0.4 | 0.71 | 0.77 | 0.85 | 0.63 |
|  | all-mpnet-base-v2 <sup>23</sup> | 0.68 | 0.44 | 0.39 | 0.28 | 0.67 | 0.78 | 0.56 | 0.54 |
|  | gte-base <sup>20</sup> | 0.67 | 0.51 | 0.44 | 0.41 | 0.69 | 0.74 | 0.77 | 0.6 |
|  | e5-base-4k <sup>24</sup> | 0.67 | 0.55 | 0.26 | 0.31 | 0.66 | 0.68 | 0.77 | 0.56 |
|  | Bio_ClinicalBERT <sup>9</sup> | 0.21 | 0.14 | 0.17 | 0.17 | 0.22 | 0.21 | 0.37 | 0.21 |
| 0.25-0.5GB | mxbai-embed-large-v1 <sup>25</sup> | 0.74 | 0.49 | 0.44 | 0.38 | 0.73 | 0.79 | 0.86 | 0.63 |
|  | UAE-Large-V1 <sup>25</sup> | 0.73 | 0.48 | 0.44 | 0.39 | 0.72 | 0.78 | 0.86 | 0.63 |
|  | GIST-large-Embedding-v0 <sup>11</sup> | 0.75 | 0.57 | 0.48 | 0.4 | 0.74 | 0.8 | 0.86 | 0.66 |
|  | bge-large-en-v1.5 <sup>19</sup> | 0.72 | 0.46 | 0.41 | 0.4 | 0.72 | 0.78 | 0.83 | 0.62 |
|  | b1ade-embed <sup>12</sup> | 0.75 | 0.53 | 0.47 | 0.42 | 0.73 | 0.8 | 0.86 | 0.65 |
| 0.5-1GB | multilingual-e5-large-instruct <sup>26</sup> | 0.75 | 0.55 | 0.32 | 0.3 | 0.64 | 0.66 | 0.83 | 0.58 |
|  | multilingual-e5-large<sup>26</sup> | 0.7 | 0.46 | 0.27 | 0.35 | 0.64 | 0.67 | 0.69 | 0.54 |
|  | Solon-embeddings-large-0.1<sup>37</sup> | 0.73 | 0.54 | 0.39 | 0.35 | 0.68 | 0.73 | 0.82 | 0.61 |
|  | bge-m3-custom-fr<sup>27</sup> | 0.76 | 0.51 | 0.31 | 0.47 | 0.66 | 0.72 | 0.73 | 0.59 |
|  | mmlw-e5-large<sup>28</sup> | 0.71 | 0.46 | 0.39 | 0.34 | 0.71 | 0.75 | 0.83 | 0.6 |
| 1-5GB | instructor-xl<sup>29</sup> | 0.73 | 0.56 | 0.42 | 0.43 | 0.72 | 0.77 | 0.79 | 0.63 |
|  | sentence-t5-xl<sup>30</sup> | 0.64 | 0.44 | 0.39 | 0.35 | 0.58 | 0.65 | 0.52 | 0.51 |
|  | sentence-t5-xxl<sup>30</sup> | 0.7 | 0.45 | 0.38 | 0.3 | 0.57 | 0.64 | 0.53 | 0.51 |
|  | gtr-t5-xxl<sup>31</sup> | 0.66 | 0.51 | 0.45 | 0.49 | 0.74 | 0.77 | 0.78 | 0.63 |
|  | SGPT-2.7B-weightedmean-msmarco-specb-bitfit<sup>32</sup> | 0.7 | 0.27 | 0.36 | 0.31 | 0.71 | 0.76 | 0.83 | 0.56 |
| >5GB | SFR-Embedding-Mistral<sup>33</sup> | 0.86 | 0.8 | 0.41 | 0.42 | 0.69 | 0.71 | 0.79 | 0.67 |
|  | e5-mistral-7b-instruct<sup>34</sup> | 0.83 | 0.76 | 0.33 | 0.36 | 0.61 | 0.59 | 0.75 | 0.6 |
|  | Linq-Embed-Mistral<sup>35</sup> | 0.86 | 0.77 | 0.41 | 0.43 | 0.7 | 0.77 | 0.75 | 0.67 |
|  | NV-Embed-v1 <sup>13</sup> | 0.35 | 0.21 | 0.48 | 0.5 | 0.77 | 0.79 | 0.84 | 0.56 |
|  | SFR-Embedding-<br>2_R <sup>36</sup> | 0.85 | 0.76 | 0.45 | 0.45 | 0.71 | 0.72 | 0.69 | 0.66 |

### Spearman Rank Correlation Across Models and Tasks

**Table 4** provides the Spearman rank correlation values for the best-performing metric of each model across the various tasks.

### MTEB Correlation

We then compared the correlations between our average task performance score (ATPS) (across all clinical and biomedical tasks), the STS score from the MTEB suite, and the overall MTEB average score **Supplementary table 2**. The STS score, which assesses model performance on tasks requiring semantic understanding akin to our studies, correlates well with the overall MTEB average. Our ATPS correlated better with the MTEB STS score (0.73) than with the MTEB average score (0.67), reflecting the role of STS as a component of the overall MTEB metric. The correlation between the STS score and MTEB score is slightly higher than ours (0.70), however, the STS score is also a component of the MTEB score, thus affecting the correlation.

### Overall Top-Ranking Models

**Table 5** ranks the models based on the Spearman values.

**Table 5:**
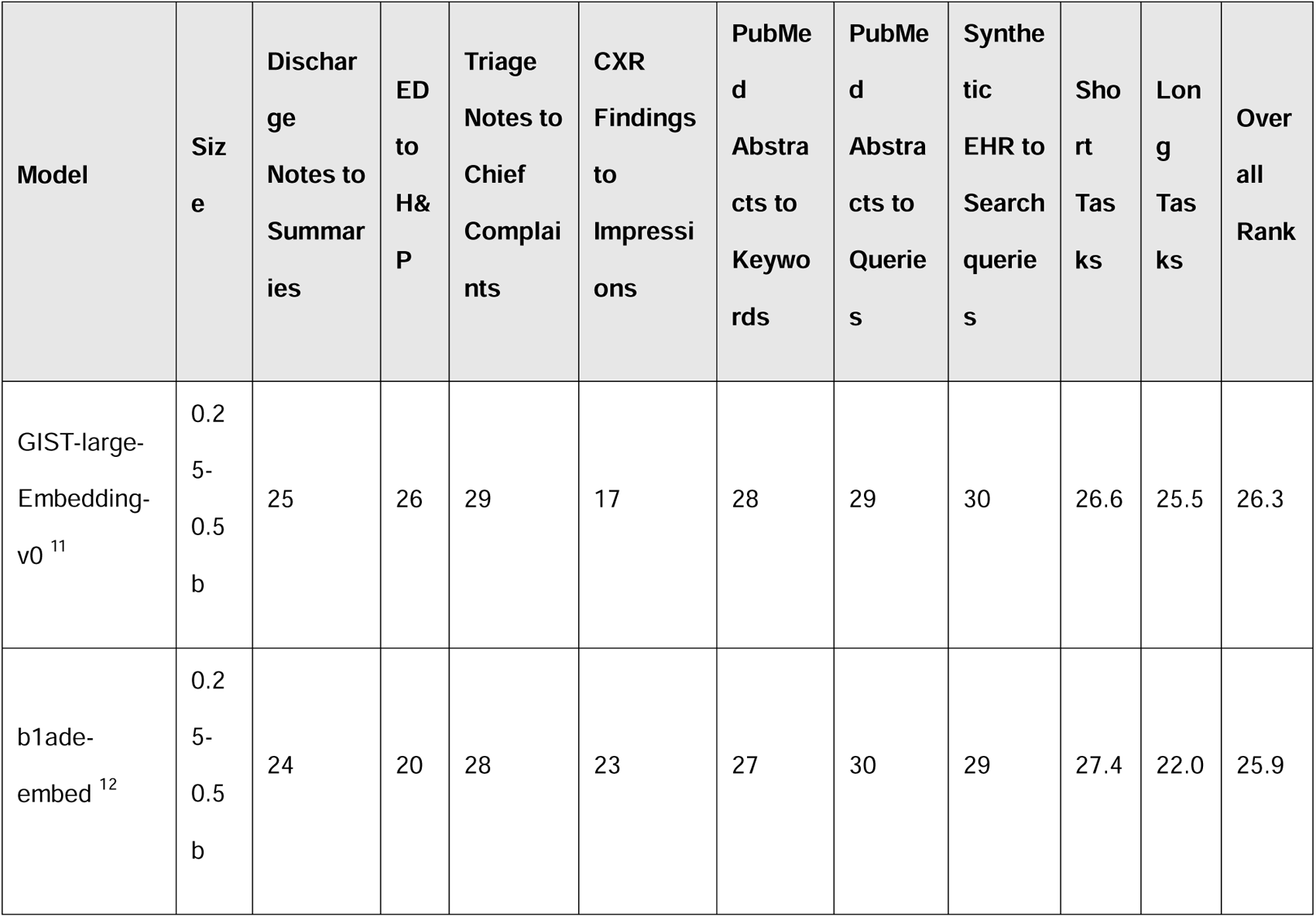
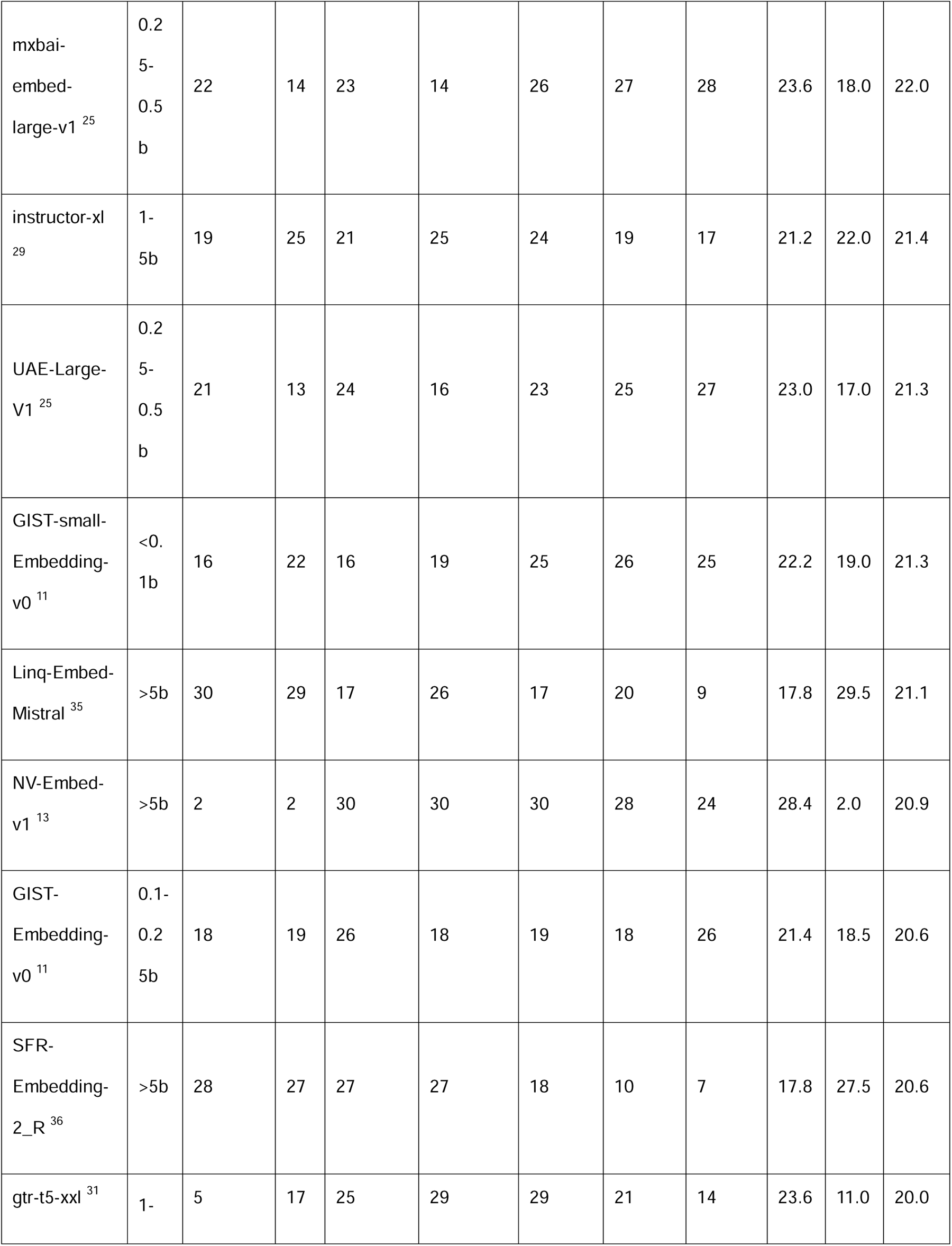
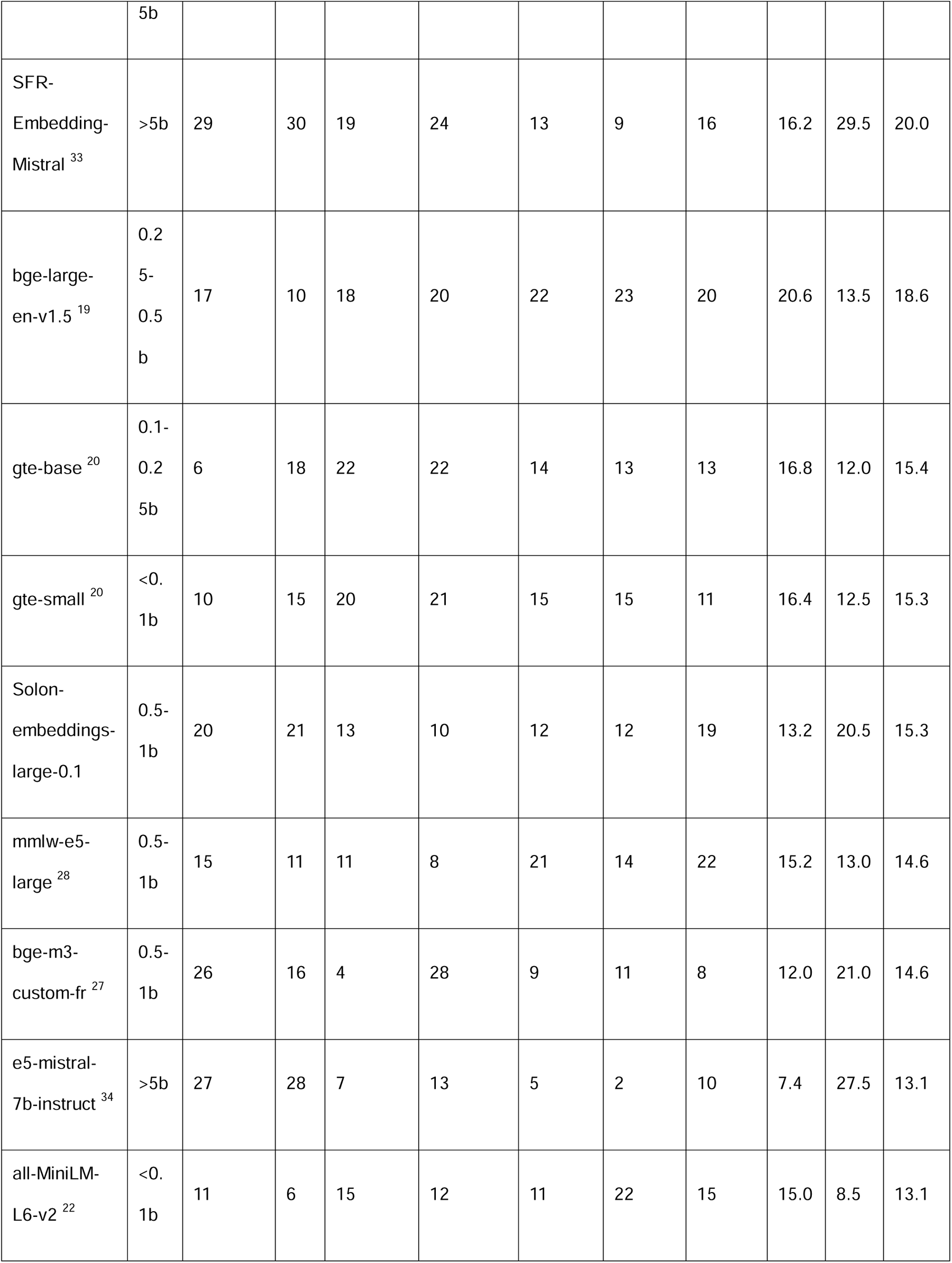
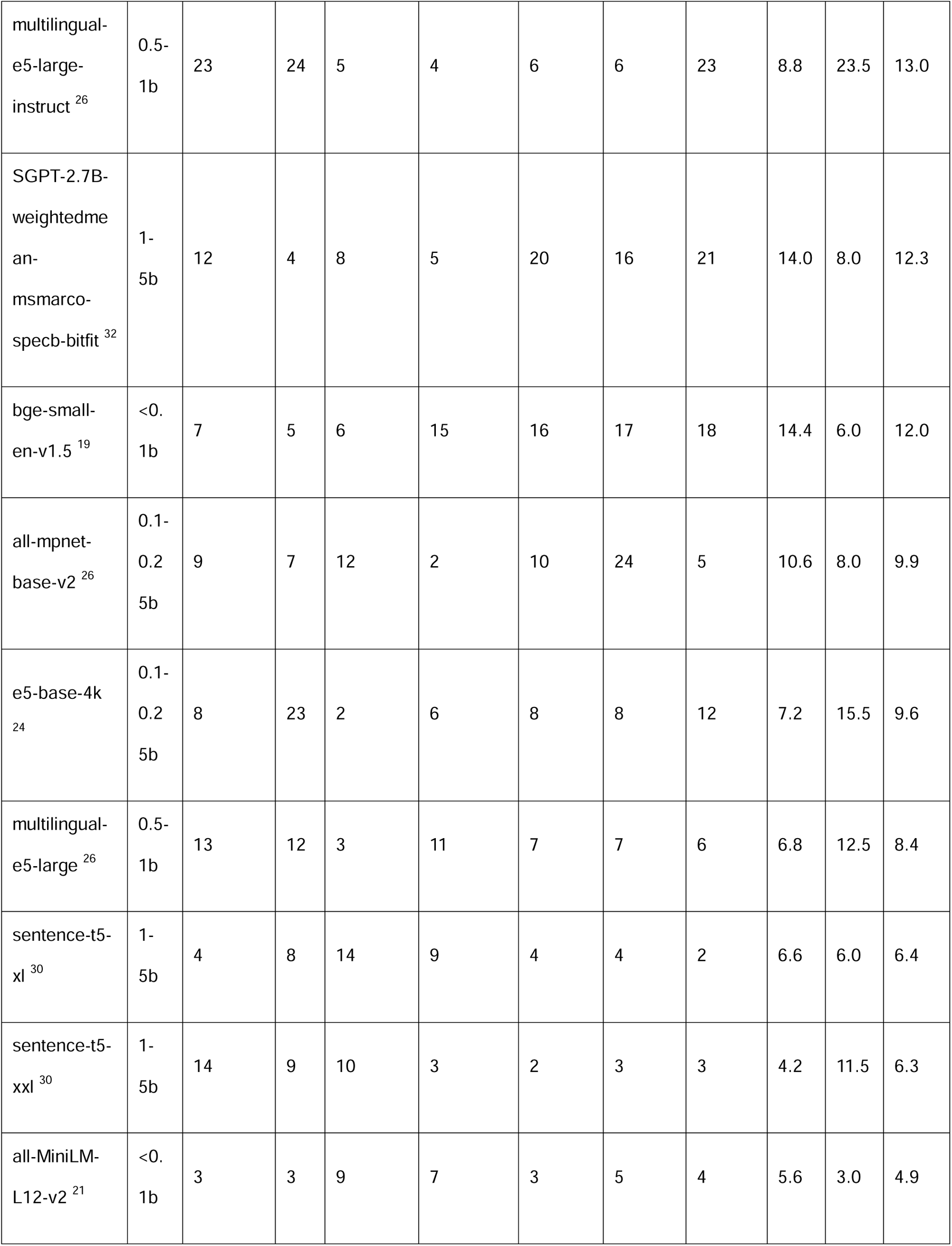
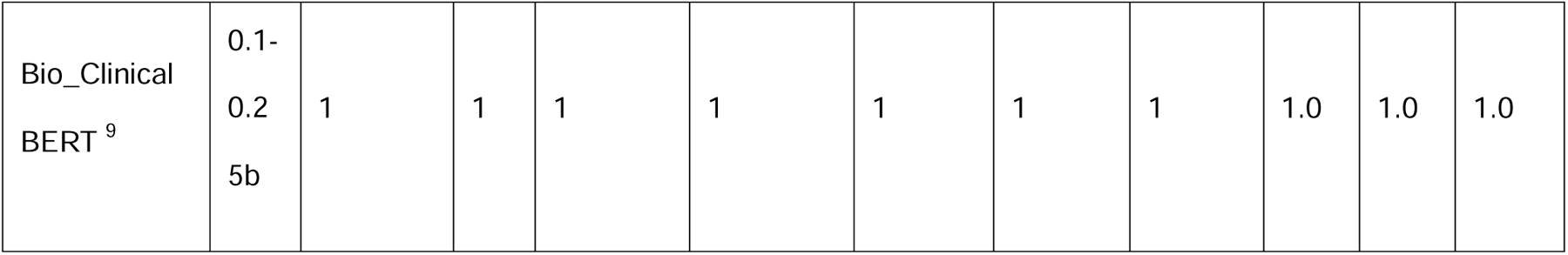
Model Rankings Across Task Types. This table outlines the rankings of the 30 embedding models across three categories of tasks: short, long, and overall. To mitigate the impact of natural variance in Spearman correlation scores between tasks, we assigned independent rankings for each task type. The ‘Bio_ClinicalBERT’ model was used as a benchmark, with a baseline rank of one in all task categories. The rankings of other models were determined based on how their Spearman scores compared to this reference model. Abbreviations: ED: Emergency Department, H&P: History & Physical Notes, CXR: Chest X-ray, EHR: Electronic Health Record

In this evaluation, the overall top-ranking models, ‘GIST-large-Embedding-v0’ and ‘b1ade-embed’ , have achieved the highest rankings across all tasks.

The ‘Bio_ClinicalBERT’ model served as a reference and recorded the lowest scores . This outcome is not surprising, as the model was not trained for semantic embedding tasks. Unlike models specifically designed for semantic embedding tasks, Bio-Clinical-BERT is essentially BERT fine-tuned on some bio-medical data.

Interestingly, the large-scale models (>5b parameters) based on Mistral-7b did not reach the top overall ranks despite their capacity.

The relationship between the overall models’ performance and their embedding times is visually represented in **Figure 1**.

**Figure 1:**
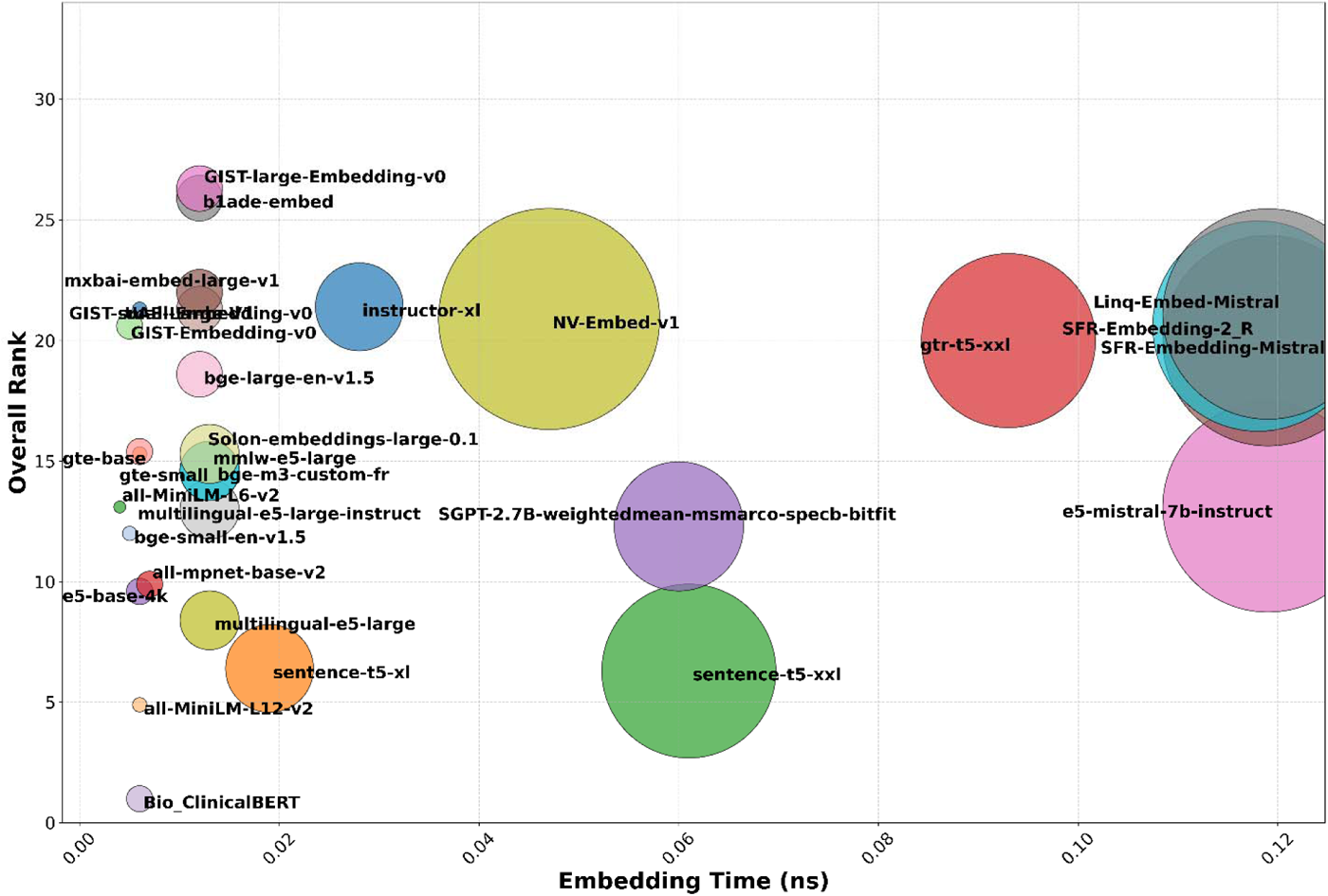
Bubble chart illustrating the relationship between the embedding models’ overall rank and their embedding time in nanoseconds (ns). Each bubble represents a different embedding model, with the size of the bubble corresponding to the model’s size. ‘GIST-large-Embedding-v0’ and ‘b1ade-embed’, have achieved the highest rankings across all tasks with low embedding times.

### Short-Tasks Top-Ranking Models

In the evaluation of short tasks (**Table 5**), ‘b1ade-embed’ and ‘GIST-large-Embedding-v0’, maintained their high performance with scores of 27.4 and 26.6, respectively. Additionally, the ‘NV-Embed-v1’, a large model and a top performer on the MTEB leaderboard, obtained the highest score, 28.4, outperforming other large models like ‘SFR-Embedding-Mistral’ and ‘Linq-Embed-Mistral’ . The relationship between the models’ performance in short tasks and their embedding times is represented in **Figure 2**.

**Figure 2:**
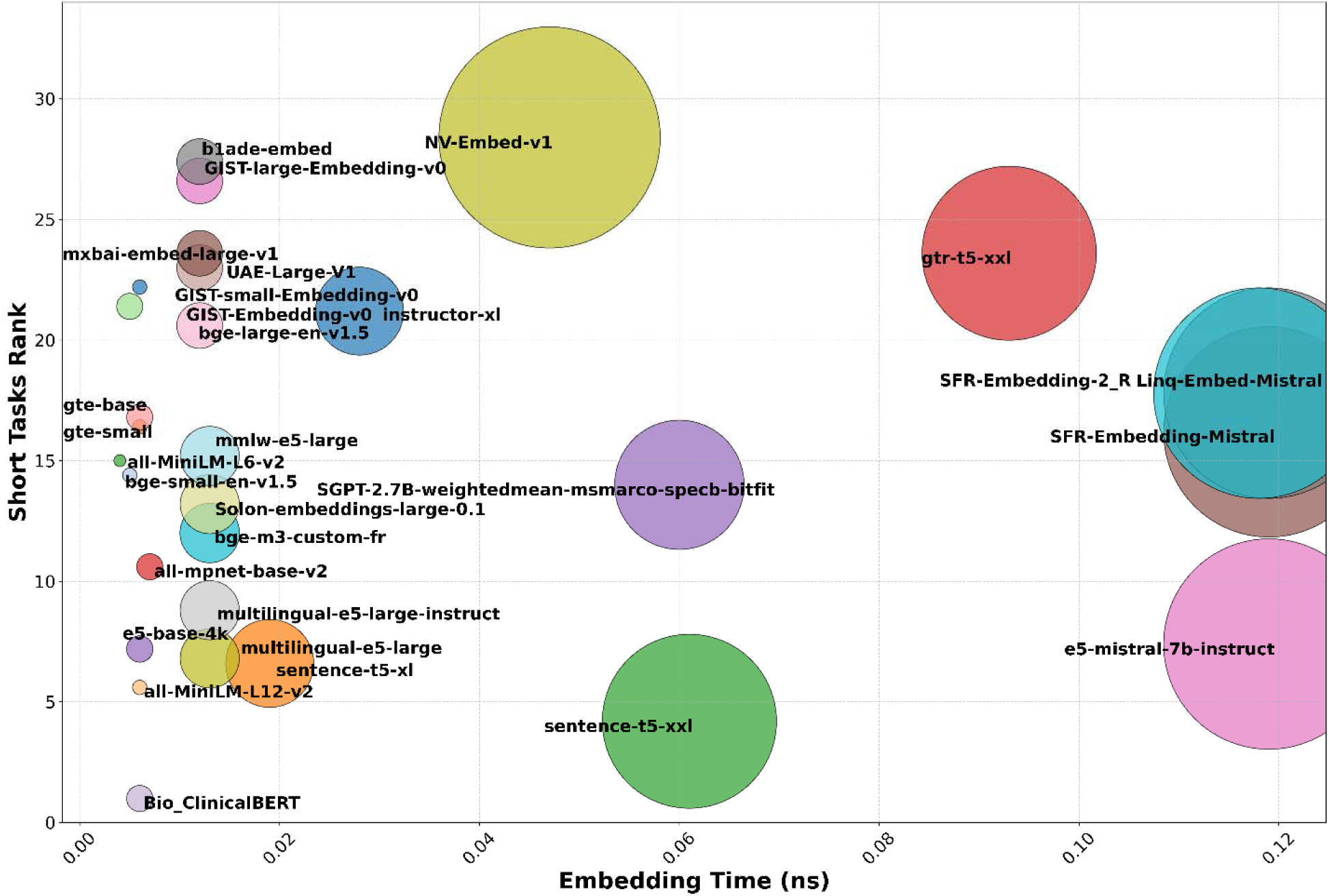
Bubble chart illustrating the relationship between the embedding models’ rankings on short tasks and their embedding time in nanoseconds (ns). Each bubble represents a different embedding model, with the size of the bubble corresponding to the model’s size. ‘NV-Embed-v1’, a large model, obtained the highest score. Models like GIST-large-Embedding-v0 and b1ade-embed show competitive rankings with relatively lower embedding times.

### Long-Tasks Top-Ranking Models

In the evaluation of long tasks (**Table** 5), the largest models with extensive context windows demonstrated dominant performance. The top four performing models in the long context tasks were >5b parameters, Mistral-7b-based models (e5-mistral-7b-instruct , SFR-Embedding-2_R , SFR-Embedding-Mistral , Linq-Embed-Mistral . Contrarily, the ‘NV-Embed-v1’, despite its large size, did not perform as well in long tasks, ranking second to last among the models evaluated, although this model was top ranging across three of the short tasks. The relationship between the models’ performance in long tasks and their embedding times is visually represented in **Figure 3**.

**Figure 3:**
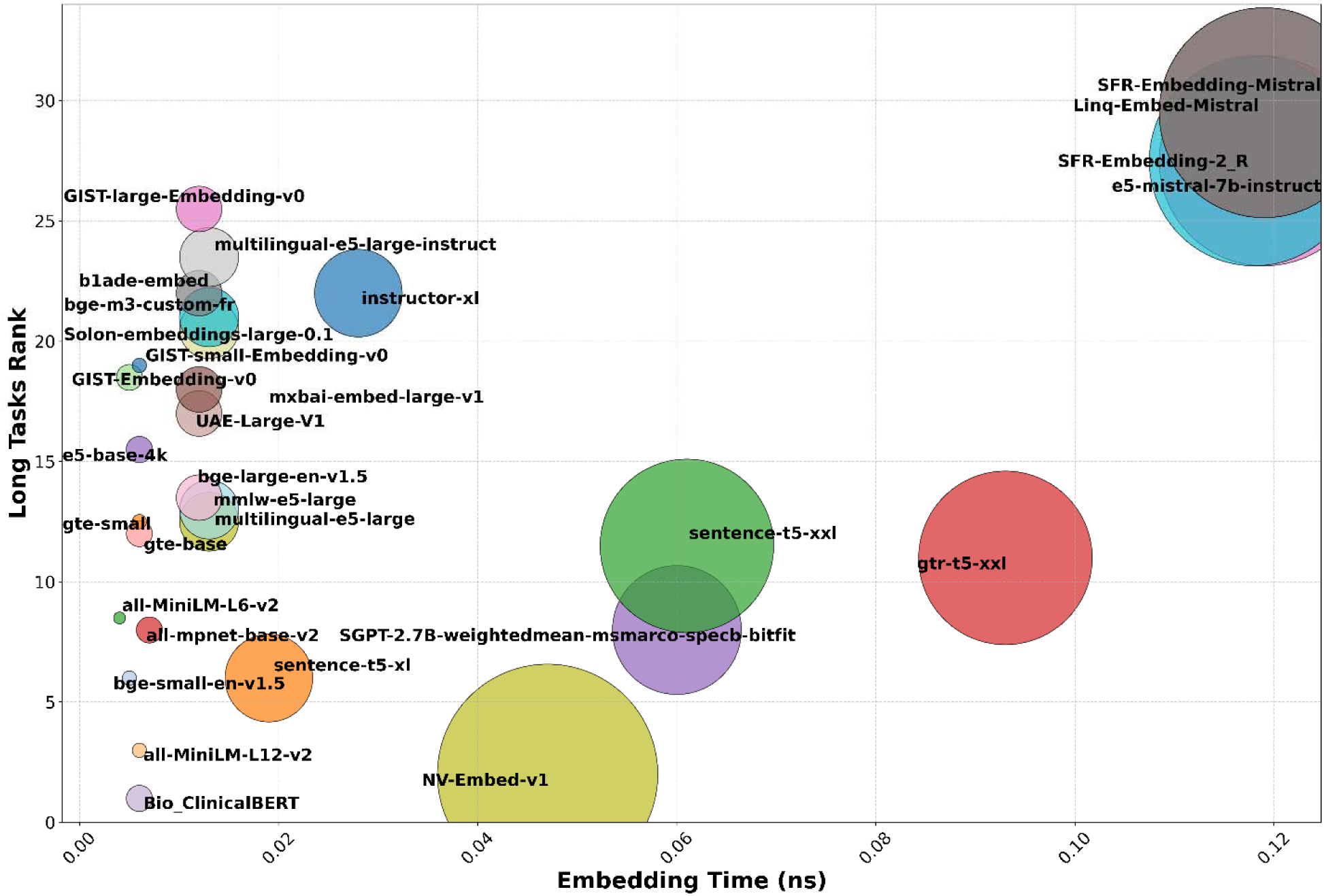
Bubble chart illustrating the relationship between the embedding models’ rankings on long tasks and their embedding time in nanoseconds (ns). Each bubble represents a different embedding model, with the size of the bubble corresponding to the model’s size. Models such as *SFR-Embedding-Mistral* and *Linq-Embed-Mistral* demonstrate strong performance with top rankings on long tasks.

### Comparison of Models Across Short Clinical vs. PubMed Tasks

In our analysis of model performance across “Short Clinical Tasks” and “Short PubMed Tasks,” (**Table** 5) ‘NV-Embed-v1’ stands out by ranking highest in both categories. Other models exhibiting strong performance in both domains include ‘b1ade-embed’ . Similarly, ‘Linq-Embed-Mistral’ and ‘instructor-xl’ show consistency, ranking in the top tiers for both clinical and PubMed tasks.

Conversely, ‘UAE-Large-V1’ performs significantly better in clinical tasks than in PubMed, whereas ‘gtr-t5-xxl’ shows a reversed trend, highlighting their specialized strengths in respective areas.

## Discussion

This study provides a framework to assess the performance of leading text embedding models for specific medical data. Additionally, it offers valuable insights into performance on specific tasks, including short and long texts, as well as biomedical and clinical data. Our findings can help identify the most effective models for those working with medical data, providing guidance for choosing models which may best suit their tasks. Our findings reveal that text embedding models, initially benchmarked on general-domain datasets, maintain high efficacy in the medical domain. The top-performing models demonstrated robust capabilities in handling medical language and contextual nuances, evidenced by their high Spearman rank correlations across various tasks.

Models like ‘GIST-large-Embedding-v0’^11^ and ‘b1ade-embed’ ^12^ excel in both short and long tasks, demonstrating superior embedding capabilities and impressive performance despite their smaller size, making them highly efficient choices.

Large-scale models, particularly those based on Mistral-7b, demonstrated superior performance in long-context tasks. This can be attributed to their longer context windows, which allow for better handling of detailed clinical texts, while shorter context window models are truncated after 512 tokens. Conversely, ‘NV-Embed-v1’^13^, despite being a large 7b model and the top performer in short tasks, did not perform as well in long tasks, indicating a potential limitation in its training for handling extended contexts. This outcome suggests that sheer model size does not guarantee superior performance across varied tasks. The adaptability and training specificity seem to play more critical roles in determining model effectiveness across such diverse testing scenarios.

The comparison between clinical and PubMed tasks revealed that certain models have specialized strengths. For instance, ‘UAE Large V1’ performed better in clinical tasks, while ‘sentence transformers gtr t5 xxl’ excelled in PubMed tasks. This differential performance emphasizes the importance of choosing models based on the specific demands of the text type and context within medical data processing. Some models are versatile across domains, while others are tailored to specific types of medical text, providing insights into their application-specific efficacy.

We address limitations specific to healthcare in the MTEB framework. First, in the MTEB framework, the statistical measure employed assesses how well the model’s outputs (similarity scores) align with actual human-assigned similarity scores ^4^. In our study, we calculated the Spearman rank correlation between metrics and data integrity levels. This provides a measure of model performance stability across different data conditions and tasks. The consistency in performance across varying levels of data integrity demonstrates the models’ ability to maintain semantic integrity. This is particularly crucial in medical applications, where data often varies in completeness and clarity.

Second, the MTEB framework does not predominantly focus on the medical domain. Within its leaderboard challenge, only a small number of biomedical datasets, such as BIOSSES ^7^ (Biomedical Sentence Similarity Estimation System) are included. BIOSSES, consisting of 100 sentence pairs annotated with semantic similarity scores by domain experts, may not capture the full diversity of biomedical literature. In contrast, our study employed a methodology that did not rely on human annotators, allowing us to utilize relatively large datasets varied in size and scope. Our method can provide an easy way to expand the types and sizes of the benchmarking datasets.

Third, the MTEB scores might reflect potential overfitting, as models’ training data could inadvertently include the datasets used in MTEB, allowing for over-fitting on these specific tasks ^14^. In contrast, our study employed a diverse set of real-world new medical tasks and datasets, providing a more accurate measure of a model’s performance in practical medical scenarios. Moreover, using our method, new benchmarking random datasets can be created dynamically. This approach ensures that the models are assessed based on their ability to handle a variety of medical data, rather than their performance on published benchmark tasks.

Another platform, “Papers with Code,” tracks the “State-of-the-Art” (SOTA) in machine learning across a range of tasks, including specific leaderboards for medical data. However, for sentence embeddings in biomedical contexts, it also just utilizes the BIOSSES benchmark, with the above-mentioned limitations ^15^.

In some studies, researchers have independently developed models tailored to text embedding within medical contexts, such as BioSentVec ^16–18^. However, these individual efforts, while valuable, do not establish benchmarks against which to measure the performance of a broad array of existing models across varied medical tasks.

Our results provide a practical framework for medical professionals and researchers working with medical data and seeking to leverage NLP technologies effectively. By identifying the optimal models for various medical tasks, we offer guidance on which models are best suited for medical applications. Additionally, several of the tasks and associated data evaluated in this study will now be available for public use, facilitating further model validation.

Our study has several limitations. Firstly, the scope of evaluated models was restricted to those available on the Hugging Face platform with implementations via the Sentence Transformers library, which may exclude potentially effective models do not present on this platform. Secondly, the generated synthetic data, particularly from the Llama-3-70b model, may not fully capture the complexity and variability of real patient data, however we did use real patient data from several databases. Additionally, our study primarily focused on English language texts, and the performance of these models on non-English medical texts remains to be explored. Furthermore, while data integrity levels were considered, the specific nature of the noise introduced may not fully capture the complexities and variations found in real-world medical data. Different types of noise (e.g., typographical errors vs. semantic errors) might affect model performance differently.

In conclusion, the suggested framework provides guidance for selecting embedding models tailored to various medical tasks. By leveraging task-specific models, we can enhance key applications such as semantic search and RAG, which, despite their potential, are still underutilized in healthcare.

## Supporting information

Supplementary Excel eTable 1

Supplementary eFigure 1

## Data Availability

All data produced in the present study are available upon reasonable request to the authors

