## Supplementary eFigure 1 for "A Scalable Framework for Benchmarking Embedding Models for Semantic Medical Tasks"

**Supplementary Content**

eFigure 1. The prompt for search query generation from PubMed abstracts....................................2

eFigure 2. Terms for Electronic Health Record (EHR) note generation..........................................3

eFigure 3. The prompt for simulating EHR notes and generating search queries ............................4

eFigure 4. The prompt for summarizing discharge notes from the MIMIC-IV database.................5

eTable 1: Common phrases indicating normal findings in Chest X-Ray reports..........................6

eTable 2: Correlation comparisons between task scores and MTEB metrics................................7

**eFigure 1: The complete prompt used** **for search query generation from PubMed abstracts, including JSON directives and formatting.**

PubMed Query:

prompt = f"""

Given the following PubMed abstract, please create a free-text search query that would typically be used to retrieve similar types of abstracts.

The free-text query should encapsulate one-three elements in the abstract to simulate a common query used in a Retrieve and Generate (RAG) system.

\#### Abstract:

{abstract}

Return in JSON format:

{{"query": "<a free-text search query that encapsulates 1-3 key elements in the abstract>"}}

Only return the JSON and not any other text, as I'm structuring your reply!!

"""

**eFigure 2.** **Terms for Electronic Health Record (EHR) note generation**

Question-Answer:

listA = ["acute myocardial infarction", "asthma exacerbation", "chronic obstructive pulmonary disease (COPD)", "autoimmune thyroiditis", "osteoarthritis flare-up", "bipolar disorder", "pancreatitis", "acute renal failure", "pneumonia", "stroke", "hypertension", "diabetes mellitus", "chronic kidney disease", "migraine", "heart failure", "arthritis", "depression", "allergic rhinitis", "GERD", "anemia", "sepsis", "multiple sclerosis", "peptic ulcer disease", "peripheral artery disease", "epilepsy", "cholecystitis", "hyperthyroidism", "hypothyroidism", "hepatitis C", "rheumatoid arthritis", "systemic lupus erythematosus", "celiac disease", "endometriosis", "ulcerative colitis", "fibromyalgia", "polycystic ovary syndrome", "sickle cell anemia", "glaucoma", "macular degeneration", "chronic fatigue syndrome"]

listB = ["undergoing an echocardiogram", "initiating insulin therapy", "adjusting antidepressant medications", "scheduling for joint replacement surgery", "prescribing a new asthma inhaler", "emergency appendectomy", "physical rehabilitation post-stroke", "starting dialysis", "antibiotic treatment for bacterial infection", "cognitive behavioral therapy sessions", "blood transfusion", "immunotherapy", "chemotherapy", "radiotherapy", "dietary modifications", "exercise regimen", "pacemaker implantation", "stent placement", "liver function tests", "colonoscopy", "CT scan", "MRI scan", "cardiac catheterization", "kidney transplant", "bone marrow biopsy", "ultrasound", "endoscopy", "laparoscopy",

"coronary artery bypass graft", "ventilator support", "pain management program", "insulin pump therapy", "speech therapy", "occupational therapy", "oxygen therapy", "proton therapy", "stem cell therapy", "gene therapy", "hormone replacement therapy", "electroconvulsive therapy”]

listC = ["following a routine check-up", "after visiting the ER with severe symptoms", "during a management review for chronic condition", "post-operative recovery period", "adjusting to a new treatment regimen", "showing symptoms despite ongoing treatment", "responding well to the new medication", "experiencing side effects from treatment", "requiring further diagnostic testing", "scheduled for a follow-up consultation", "after a recent hospital discharge", "in the midst of a clinical trial", "as part of a regular monitoring program", "during a flare-up of symptoms", "after a recent infection", "due to a sudden onset of symptoms", "following a specialist referral", "during a telemedicine consultation", "following a lifestyle change", "after a family history assessment", "following a new diagnosis", "after a sudden weight loss", "due to recurring symptoms", "after a recent surgery", "as part of a treatment evaluation", "following an allergic reaction", "due to persistent pain", "after experiencing dizziness", "during a routine vaccination visit", "after a screening test result", "as part of a preoperative assessment", "due to unexpected test results", "following a mental health evaluation", "after a fall or injury", "during a routine cancer screening", "after a cardiovascular event", "due to ongoing gastrointestinal issues", "after being referred by another physician", "following a blood pressure check", "during a post-discharge follow-up"]

subjectA = random.choice(listA)

subjectB = random.choice(listB)

subjectC = random.choice(listC)

**eFigure 3. The complete font of the prompt** **used for simulating Electronic Health Record (HER) notes and generating search queries with the JSON directive and formatting**

prompt = f"""

Please generate a detailed paragraph of approximately 150 words that simulates an excerpt from an EHR note.

The note should involve the following contents: {subjectA}, {subjectB}, {subjectC}.

Ensure the paragraph reflects realistic medical documentation with a coherent narrative flow.

Return in JSON format:

{{"paragraph":<your 150-word medical content paragraph>}}"""

prompt = f"""

Given the following EHR note paragraph, please create a search query that would typically be used to retrieve this type of record in a medical information system.

The query should encapsulate one-three key elements and medical terms in the paragraph to ensure precision in a Retrieve and Generate (RAG) system.

For instance, if a note details a "45-year-old female diagnosed with early-stage breast cancer, undergoing lumpectomy followed by radiation therapy, and experiencing side effects such as mild fatigue and skin irritation,"

the corresponding search query might be: "Retrieve cases of early breast cancer patients undergoing procedures."

\#### EHR Note Paragraph:

{paragraph}

Return in JSON format:

{{"query": "<a search query that encapsulates the key elements and medical terms in the paragraph>"}}

Only return the JSON and not any other text, as I'm structuring your reply!!"""

**eFigure 4. The complete font of the prompt used for summarizing discharge notes from the MIMIC-IV database with the JSON directive and formatting**

Llama summarizations:

“““Please provide a short summary of the following patient discharge note.

Return your answer in a json format:

{{"summary":<your summary>}}”””

**eTable 1: A list of phrases used to categorize “normal” findings in the MIMIC IV Chest X-Ray Reports**

**Common Phrases Indicating Normal Chest X-Ray Findings**

"No acute cardiopulmonary process”

"No acute intrathoracic process"

"No acute cardiopulmonary abnormality"

"No evidence of acute cardiopulmonary process"

"No new focal opacities to suggest pneumonia"

"Stable chest radiograph"

"No acute cardiothoracic process"

**eTable 2: Correlation Comparisons Between Task Scores and MTEB Metrics.** This table compares the correlations between the average task performance score across all tasks and two key metrics from the MTEB suite: the STS score and the overall MTEB average score.

Abbreviations: MTEB: Medical Text Embedding Benchmark, STS: Semantic Textual Similarity: STS

| **Model** | **Model Size Group** | **Average Task Performance Score (ATPS)** | **Mean STS Score** | **Mean MTEB Score** |
| --- | --- | --- | --- | --- |
| GIST-small-Embedding-v0 ^11^ | <0.1b | 0.63 | 83.03 | 62.72 |
| bge-small-en-v1.5 ^19^ | <0.1b | 0.57 | 81.59 | 62.17 |
| gte-small ^20^ | <0.1b | 0.6 | 82.07 | 61.36 |
| all-MiniLM-L12-v2 ^21^{, 2021 #3776} | <0.1b | 0.48 | 79.8 | 56.53 |
| all-MiniLM-L6-v2 ^22^ | <0.1b | 0.58 | 78.9 | 56.26 |
| GIST-Embedding-v0 ^11^ | 0.1-0.25b | 0.63 | 83.51 | 63.71 |
| all-mpnet-base-v2 ^23^ | 0.1-0.25b | 0.54 | 80.28 | 57.78 |
| gte-base ^20^ | 0.1-0.25b | 0.6 | 82.3 | 62.39 |
| e5-base-4k ^24^ | 0.1-0.25b | 0.56 | 81.05 | 61.5 |
| Bio_ClinicalBERT ^9^ | 0.1-0.25b | 0.21 | nan | nan |
| mxbai-embed-large-v1 ^25^ | 0.25-0.5b | 0.63 | 84.9 | 63.25 |
| UAE-Large-V1 ^25^ | 0.25-0.5b | 0.63 | 84.54 | 64.64 |
| GIST-large-Embedding-v0 ^11^ | 0.25-0.5b | 0.66 | 84.59 | 64.34 |
| bge-large-en-v1.5 ^19^ | 0.25-0.5b | 0.62 | 83.11 | 64.23 |
| b1ade-embed ^12^ | 0.25-0.5b | 0.65 | 85.04 | 64.21 |
| multilingual-e5-large-instruct ^26^ | 0.5-1b | 0.58 | 84.78 | 64.41 |
| multilingual-e5-large ^26^ | 0.5-1b | 0.54 | 82.06 | 61.42 |
| Solon-embeddings-large-0.1 | 0.5-1b | 0.61 | nan | nan |
| bge-m3-custom-fr ^27^ | 0.5-1b | 0.59 | nan | nan |
| mmlw-e5-large ^28^ | 0.5-1b | 0.6 | nan | nan |
| instructor-xl ^29^ | 1-5b | 0.63 | 83.06 | 61.79 |
| sentence-t5-xl ^30^ | 1-5b | 0.51 | 81.66 | 57.87 |
| sentence-t5-xxl ^30^ | 1-5b | 0.51 | 82.63 | 59.51 |
| gtr-t5-xxl ^31^ | 1-5b | 0.63 | 78.38 | 58.97 |
| SGPT-2.7B-weightedmean-msmarco-specb-bitfit ^32^ | 1-5b | 0.56 | 76.83 | 57.15 |
| SFR-Embedding-Mistral ^33^ | >5b | 0.67 | 85.05 | 67.56 |
| e5-mistral-7b-instruct ^34^ | >5b | 0.6 | 84.63 | 66.63 |
| Linq-Embed-Mistral ^35^ | >5b | 0.67 | 84.97 | 68.17 |
| NV-Embed-v1 ^13^ | >5b | 0.56 | 82.84 | 69.32 |
| SFR-Embedding-2_R ^36^ | >5b | 0.66 | 81.26 | 70.31 |
